# Evaluating the association between household water, sanitation and hygiene (WASH) and selected placenta-related complications in The Gambia, Kenya and Mozambique

**DOI:** 10.1101/2025.08.13.25332637

**Authors:** Joseph Wells, Joseph Waiswa, Anifa Vala, Bahtisen Golla, Grace Mwashigadi, Hawanatu Jah, Marleen Temmerman, Robin Okello, Isaac Mwaniki, Herminio Cossa, Moses Mukhanya, Esperança Sevene, Yahaya Idris, Onesmus Wanje, Angela Koech, Anna Roca, Laura Braun, Marie-Laure Volvert, Fatima Touray, Umberto D’Alessandro, Peter von Dadelszen, Wendy Graham, Hannah Blencowe, the PRECISE and PRECISE-DYAD Networks (Table S1 and S2)

## Abstract

Placenta-related complications such as pre-eclampsia, small-for-gestational-age (SGA) and stillbirth contribute significantly to maternal and perinatal mortality globally, with the highest burden in low– and middle-income countries (LMICs). The potential impact of inadequate access to household water, sanitation, and hygiene (WASH) on these outcomes has not been quantified. This study investigates the association between household WASH conditions and pre-eclampsia, stillbirth and SGA in The Gambia, Kenya, and Mozambique, where access to safe WASH services remains a challenge.

This study is nested within the PRECISE (PREgnancy Care Integrating Translational Science, Everywhere) study, a prospective observational cohort including 5,745 unselected pregnant women. Multivariate logistic regression analysis was used to test the associations between household WASH and pre-eclampsia, SGA or stillbirth. Compared to women with piped water in their homes, those relying on other improved water sources had higher odds of experiencing selected placenta-related complications (adjusted odds ratio (aOR) 1.36 [95% confidence interval (CI) 1.18, 1.57], p < 0.001). Country-specific analyses revealed differences across settings. In both The Gambia (1.52 [1.03, 2.24], p = 0.034) and Kenya (1.29 [1.04, 1.59], p = 0.019), the use of other improved water sources was associated with increased odds of selected placenta-related complications. Unimproved sanitation, compared with improved sanitation, was associated with increased odds of selected placenta-related complications in Mozambique (1.35 [1.02, 1.80], p = 0.038).

The findings highlight the role of household-level WASH conditions as potential risk factors for placenta-related complications. Even when water sources are improved, contamination can occur during collection, transport and storage, while unimproved sanitation can increase pathogen exposure. These results underline the need for targeted WASH interventions to reduce pregnancy-related risks. Addressing these gaps could significantly reduce the prevalence of placenta-related complications, contributing to improved maternal and neonatal health outcomes in LMICs. Future research should explore the mechanisms linking WASH to pre-eclampsia, SGA and stillbirth and other placenta-related complications, and assess the effect of comprehensive WASH interventions.

## Introduction

Placenta-related complications, such as pre-eclampsia, fetal growth restriction, and stillbirth, contribute to approximately 46,000 maternal deaths and 2.5 million fetal, neonatal, and infant deaths globally each year (1). The majority (99%) of these deaths occur in low– and middle-income countries (LMICs), with over half taking place in sub-Saharan Africa (1). A range of factors have been associated with these complications. However, in many LMICs, the complete array of risk factors for these conditions remains poorly understood.

One pathway of interest is the potential increased risk of placenta-related complications in women exposed to inadequate household water, sanitation and hygiene (WASH) conditions. Poor household– and community-level WASH conditions are well-established determinants of various adverse health outcomes, particularly diarrhoeal diseases. Improved WASH infrastructure can help protect against diarrhoeal pathogens and reduce exposure to soil-transmitted helminths, acute respiratory infections and the risks associated with chemical contaminants in water (2–4).

Exposure to poor WASH conditions during pregnancy can have both direct effects, such as acute maternal gastrointestinal infection or exposure to chemical contaminants through drinking unsafe water, as well as indirect effects, such as increased physical exertion from water collection. These effects could in turn potentially be associated with a wide range of maternal and reproductive health complications, including maternal stress, morbidity or mortality, preterm birth, small-for-gestational-age (SGA), low birth weight (LBW), stillbirth, and neonatal deaths (4–11). However, to date, there has been limited research that specifically explores the associations between poor WASH conditions and pre-eclampsia, SGA and stillbirth.

This study examines household WASH characteristics among pregnant women in a large cohort from The Gambia, Kenya, and Mozambique, and explores their associations with selected placenta-related complications of pre-eclampsia, SGA and stillbirth (12). National data from these three countries indicate poor WASH infrastructure in households. In The Gambia, 41% of households rely on unimproved sanitation facilities, such as pit latrines without a slab or bucket latrines, or practice open defecation. In Kenya, this figure is 38%, while in Mozambique it is 58% (13). Additionally, many households in these countries use unimproved water sources, such as unprotected wells or springs, or surface water. In The Gambia, this figure is 9%, in Kenya, 28% and in Mozambique, 27% (13). By assessing whether household WASH characteristics are associated with pre-eclampsia, SGA and stillbirth, we aim to enhance the understanding of potential risk factors. These findings could inform the design, testing, and introduction of targeted interventions to improve household-level WASH access for pregnant women, potentially reducing the incidence of placenta-related complications and the consequences for mothers and newborns.

## Methods

### Study design

This study is nested within the PRECISE (PREgnancy Care Integrating Translational Science, Everywhere) prospective observational cohort, exploring selected placenta-related complications, including pre-eclampsia, SGA and stillbirth in The Gambia, Kenya and Mozambique. This was a collaboration with Kings College London, the Medical Research Council (MRC) Unit The Gambia at the London School of Hygiene and Tropical Medicine (LSHTM), the Aga Khan University in East Africa, and the Centro de Investigação de Saúde de Manhiça in Mozambique (12, 14).

Recruitment to the PRECISE cohort occurred between 18^th^ June 2019 and 2^nd^ July 2021 in The Gambia, 24^th^ June 2019 and 6^th^ December 2022 in Kenya and 2^nd^ July 2019 and 16^th^ November 2021 in Mozambique. There was a pause in recruitment (and follow-up in The Gambia and Kenya) from April – July 2020 due to the COVID-19 pandemic (12).

### Study sites

In The Gambia, the study was conducted in the district of Farafenni, Central Gambia, on the trans-Gambian road linking Dakar with Ziguinchor, in southern Senegal. The recruiting centres were Farafenni District Hospital (urban), and two rural primary health care facilities in Illiasa and Nyagen Sanjal. In Kenya, recruiting centres were Mariakani Subcounty Hospital (urban) in Kaloleni sub-county, and Rabai Subcounty Hospital (rural) in Rabai sub-county. Both hospitals are in Kilifi County in coastal Kenya. In Mozambique, recruiting centres were Manhiça District Hospital (primarily urban population) and Xinavane Rural Hospital (primarily rural population) in Maputo Province, Southern Mozambique.

### Study Population

The PRECISE prospective pregnancy cohort enrolled unselected pregnant women aged between 15 and 49 years attending the study facilities and not referred from another facility (14). Women enrolled in the PRECISE cohort with complete household WASH and pregnancy outcome data were included in this analysis (Figure 1) (12).

**Figure 1.**
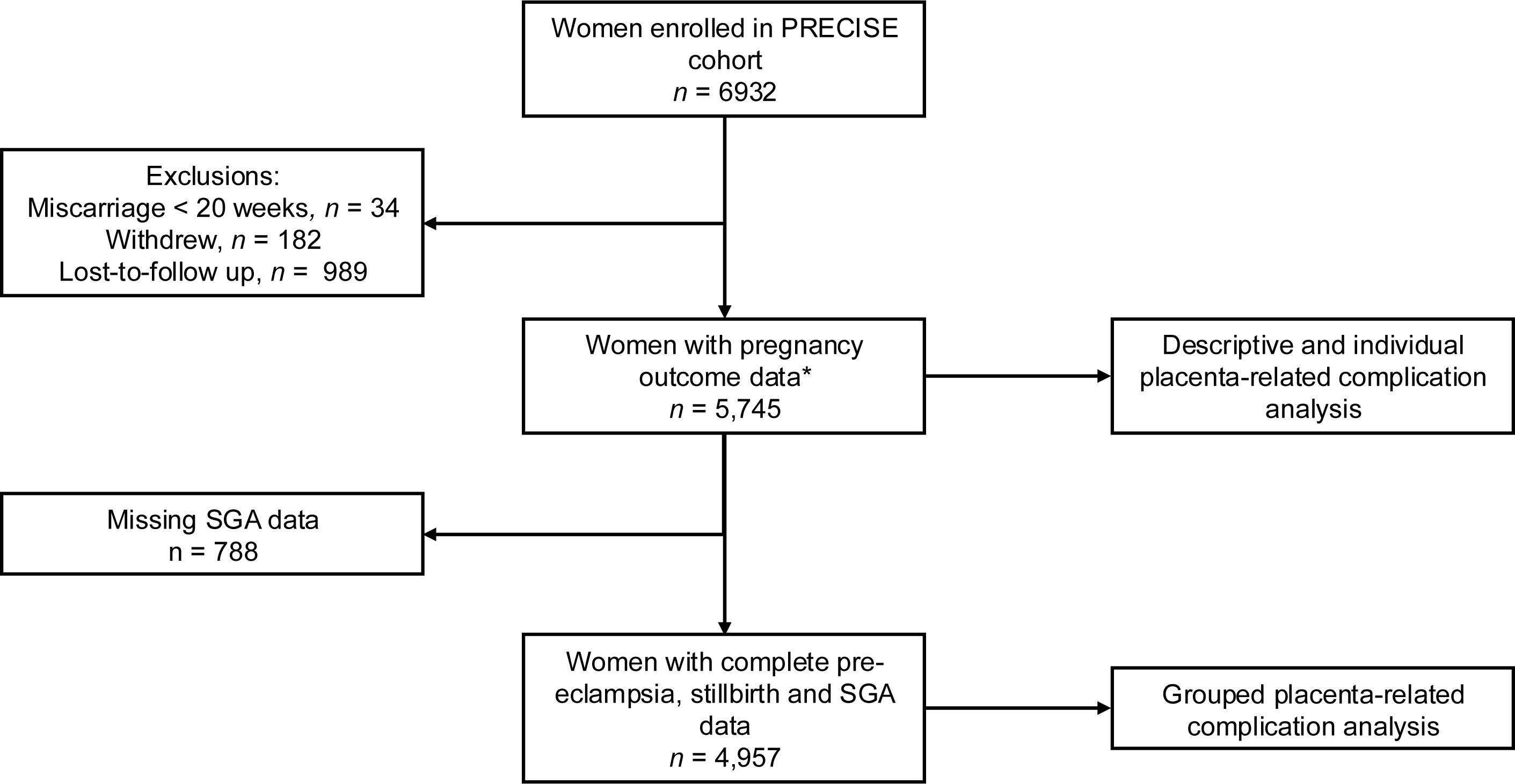
Participant flow diagram included from the PRECISE cohort. * Eighteen pregnancy outcomes could not be classified but were included in this analysis (12).

### Data collection

Enrolment took place at the first antenatal care (ANC) visit (median 20.7 weeks of pregnancy (interquartile range (IQR): 13.4, 22.6)). Information on baseline demographics, anthropometry and WASH variables were collected at enrolment. Data regarding selected placenta-related complications (pre-eclampsia, SGA or stillbirth) were collected at birth.

### Data categorisations

#### Exposure variables

Water sources and sanitation facilities were categorised into improved and unimproved using the WHO/UNICEF Joint Monitoring Programme (JMP) definitions (Figure 2) (13). Improved water sources were further subdivided into piped water into the dwelling/plot and other improved water sources. WASH practices were also categorised into the JMP service levels, used globally for the monitoring of access, availability, and quality of water, management of excreta from households, and the conditions and practices related to hygiene. These service levels were compared with the national data from the JMP global database (13).

**Figure 2.**
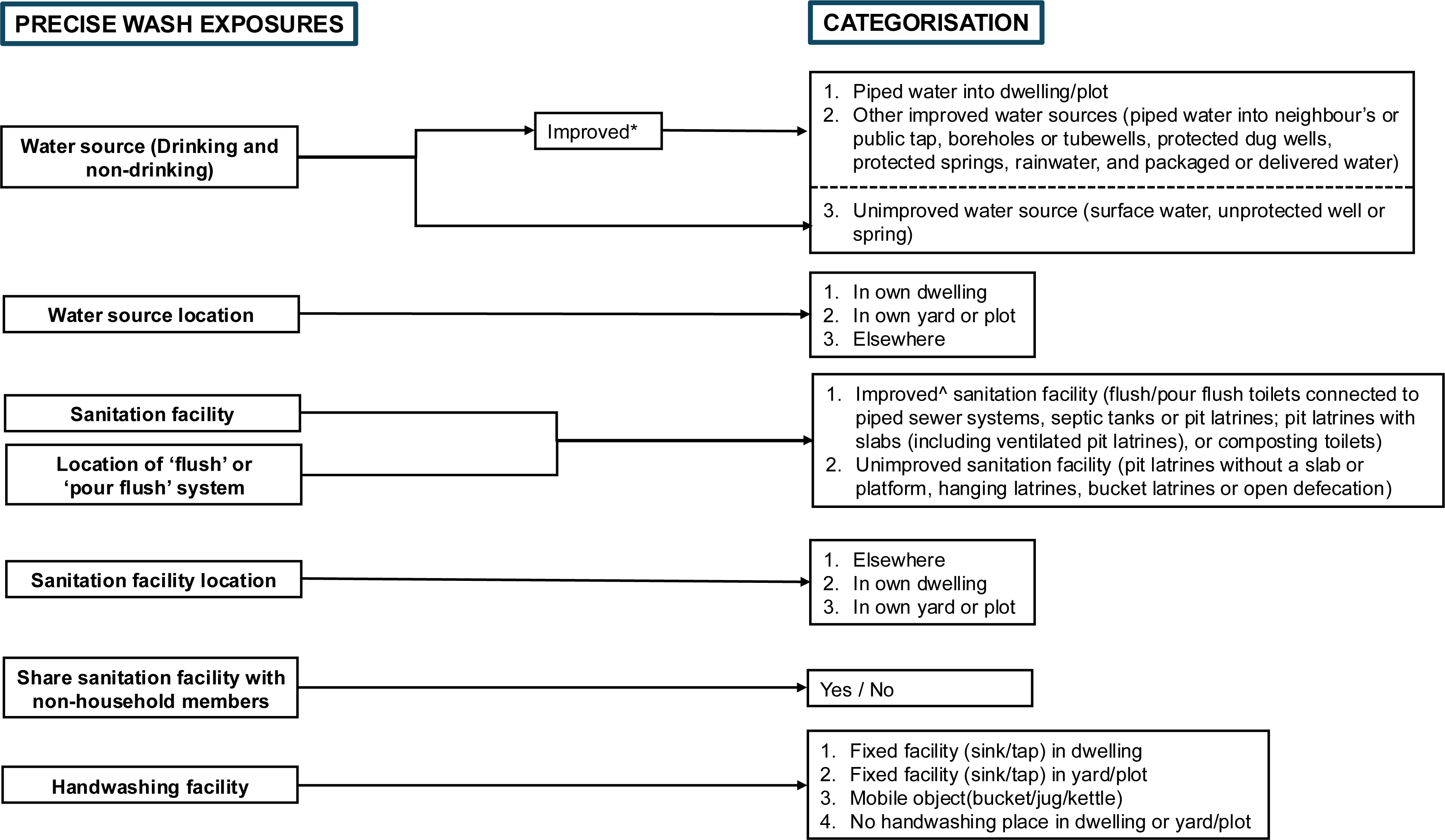
Water, sanitation and hygiene (WASH) survey questions in the PRECISE study and categorisations. * Improved drinking-water sources are defined as those more likely to be protected from outside contamination and faecal material. The improved water source variable was subdivided into piped water into dwelling/plot and “other improved sources” for further interpretability. ^Improved sanitation facilities separate human waste from human contact.

#### Selected placenta-related complications

- **Pre-eclampsia** was defined according to the International Society for the Study of Hypertension in Pregnancy criteria as: hypertension (systolic blood pressure ≥ 140mmHg or diastolic blood pressure ≥90mmHg) and at least one of:
  Symptoms: abdomen pain, headache, visual, chest pain, shortness of breath
  Signs: oxygen saturation (pulse oximetry) <90%, eclampsia, seizure, blindness, stroke, or pulmonary oedema
  Laboratory tests: thrombocytopaenia, or elevated serum creatinine, alanine transaminase, or aspartate transaminase
  Significant proteinuria (≥ ++ dipstick, urinary protein:creatinine ratio >30mg/mmol)
  Evidence of uteroplacental dysfunction: fetal death, stillbirth, placental abruption, or birthweight <5^th^ centile for gestational age (15)
- **SGA:** <10th centile for sex and gestational age, using the INTERGROWTH standards (16)
- **Stillbirth:** Baby born without vital signs at ≥20+0 weeks’ gestation or ≥500g (17)

Birth outcomes (still-or livebirth) were complete for all women, and minimal missing data (0.1%) were present for the pre-eclampsia outcome. Given the high rate of missing data (13.7%) for SGA, baseline characteristics were compared between participants with missing SGA data and those with complete data (Table S3). Due to differences between the groups, a complete case analysis was conducted for the selected placenta-related complication outcome.

### Conceptual framework

A conceptual framework was developed based on existing literature to illuminate the potential relationships between WASH and selected placenta-related complications, to inform the selection of covariates and confounders for analysis, and to support the interpretation of results (Figure 3).

**Figure 3.**
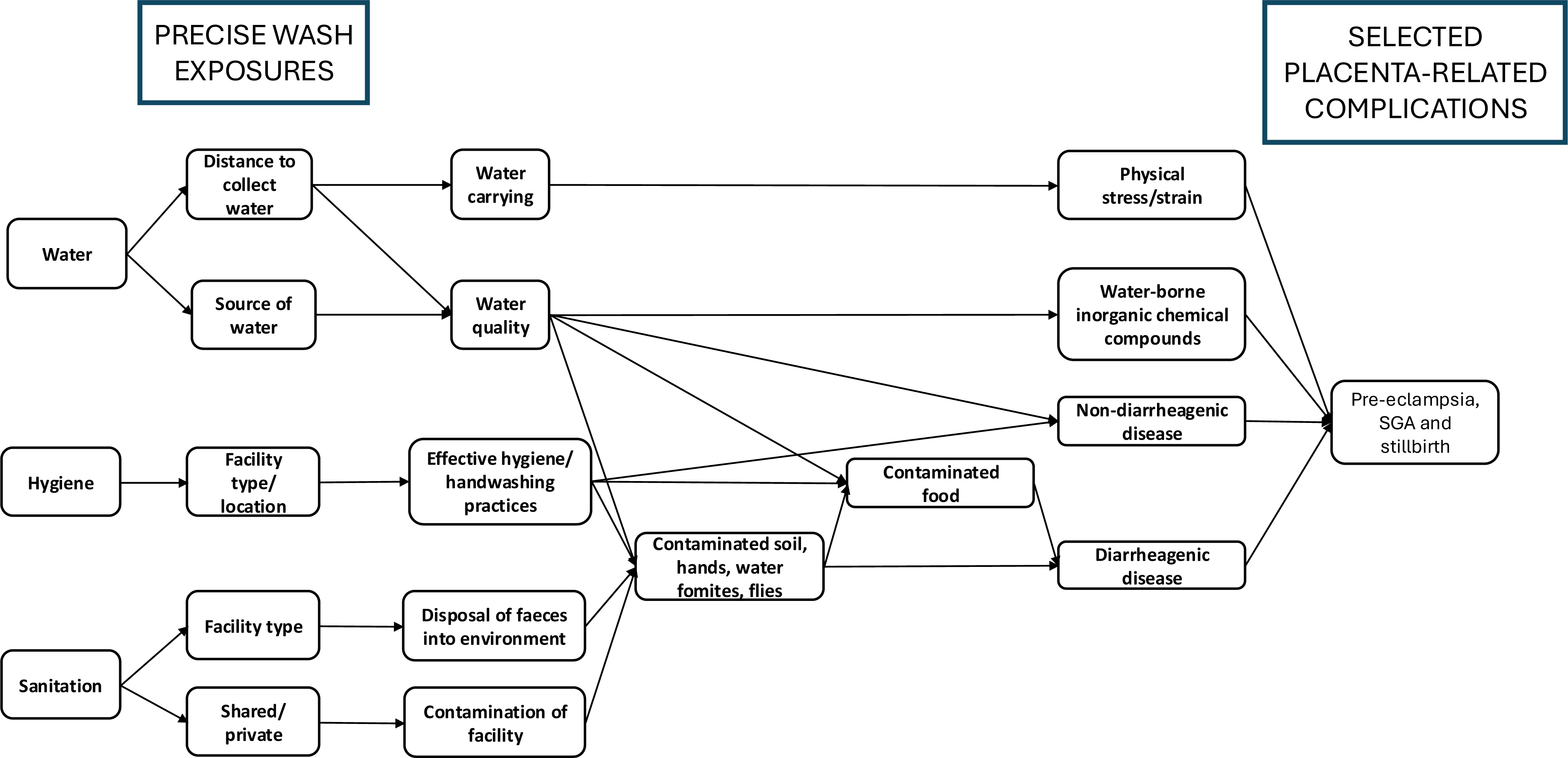
PRECISE-specific WASH exposures and potential pathways linking them to selected placenta-related complications. PRECISE, PREgnancy Care Integrating translational Science, Everywhere; SGA, small-for-gestational-age; WASH, Water, Sanitation and Hygiene.

### Potential Covariates and Confounders

Maternal and household factors, including maternal age at enrolment, parity, underweight status (defined as an average middle-upper-arm-circumference (MUAC) <23 cm), rural household location, education level, percentage likelihood of living below the poverty line and season at conception, were selected *a priori* for inclusion in the multivariate models as potential covariates and confounders. Missing covariate data were low (<5%) and were imputed using multiple imputation runs, with 20 imputed data sets and 10 iterations (mice package: ‘mice function’ (18)).

### Statistical analysis

All statistical analyses were conducted in RStudio (R Foundation for Statistical Computing) (19).

Univariate and multivariate analyses were conducted to test for associations between WASH variables and the binary selected placenta-related complication outcome. Individual variables were tested for associations using univariate binomial logistical regression (stats package: ‘glm’ function (19)).

WASH exposure variables with a p-value < 0.2 in the binomial logistical regression were considered for inclusion in the binomial portion of the multivariable model. Pearson’s correlation coefficients were generated for all combinations of selected variables with p-value <0.2. Where r^2^ > 0.5, the variable with the lowest Akaike information criterion (AIC) was selected, and the other was excluded. A stepwise model selection approach was used to identify the optimal multivariate model, with *a priori c*onfounders and covariates retained in all models (mass package: ‘stepAIC’ function (20)) (21). Variables were added in order of best model fit (AIC). An added variable was discarded where it did not improve the model fit (AIC), and the next variable was then added. This process continued until all variables had been trialled. The final multivariable model was checked for multi-collinearity, with variables exceeding a variance inflation factor of 5 removed (car package: ‘vif’ function (22)).

### Sub-group analysis

The selected multivariate variables were run with the individual country datasets. The final WASH variables were also used in a model where the outcome is changed from grouped to individual placenta-related complications: pre-eclampsia, SGA and stillbirth.

### Ethical permission statement

The PRECISE and PRECISE-DYAD studies were reviewed and approved by the King’s College London Research Ethics Committee, United Kingdom (approval references HR-17/18–7855 and HR-20/21-19714); The Gambia Government/Medical Research Council Joint Ethics Committee, The Gambia (approval references SCC 1619 and SCC 22843); the Aga Khan University Hospital Research and Ethics Committee, Nairobi, Kenya (approval references 2018/REC-74 and 2021/IERC-08); the Centro de Investigação em Saúde de Manhiça Institutional Bioethics Committee, Manhiça, Mozambique (approval reference CCI/044/OUT/2024); and the Mozambique Ministry of Health National Bioethics Committee for Health (Comité Nacional de Bioética para a Saúde, CNBS), Maputo, Mozambique (approval references 545/CNBS/18 and 655/CNBS/20). Ethical approval was granted by all committees. Written informed consent was obtained from all participants prior to their enrolment in the study.

## Results

### Characteristics of study participants

A total of 5,745 participants were included in this study, 1,178 from The Gambia, 2,706 from Kenya and 1,861 from Mozambique (Table 1). The median age of participants was 26 years (IQR: 21 – 31). Parity was most commonly between 2 and 4 births in The Gambia (41%) and Kenya (38%), but in Mozambique nulliparity (0) was more frequent (39%). The median BMI, measured at the booking appointment, was 23.8 (IQR: 21.3 – 27.1) and the median MUAC was 26.8 cm (IQR: 24.5 – 29.5) (Table 1). Overall, 10% of women were underweight (MUAC < 23 cm), with the highest prevalence in Kenya (14%) and the lowest in Mozambique (3%).

**Table 1.**
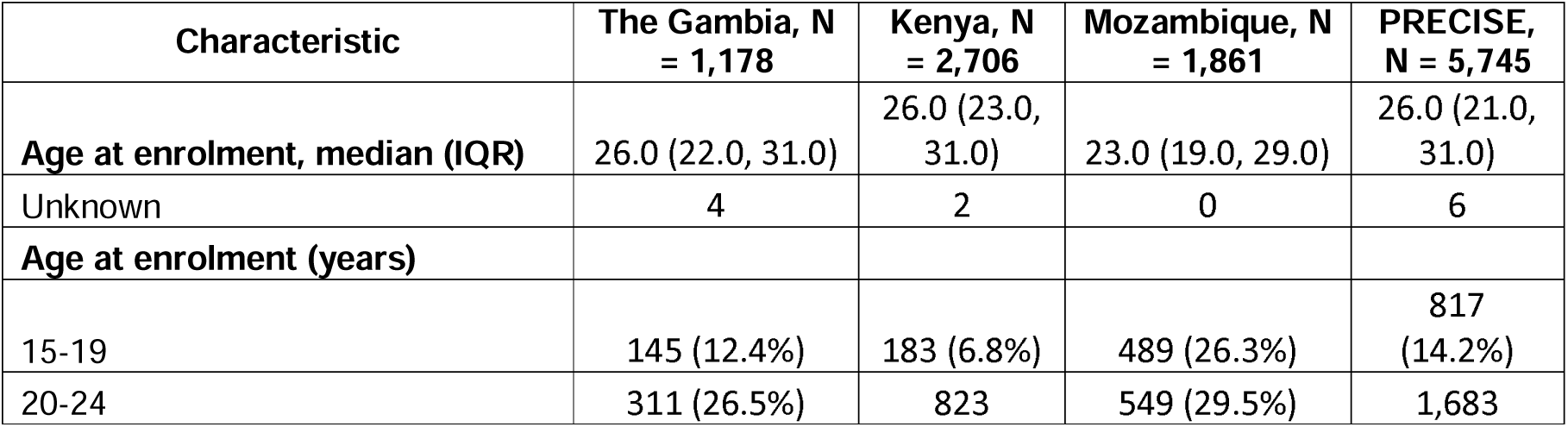
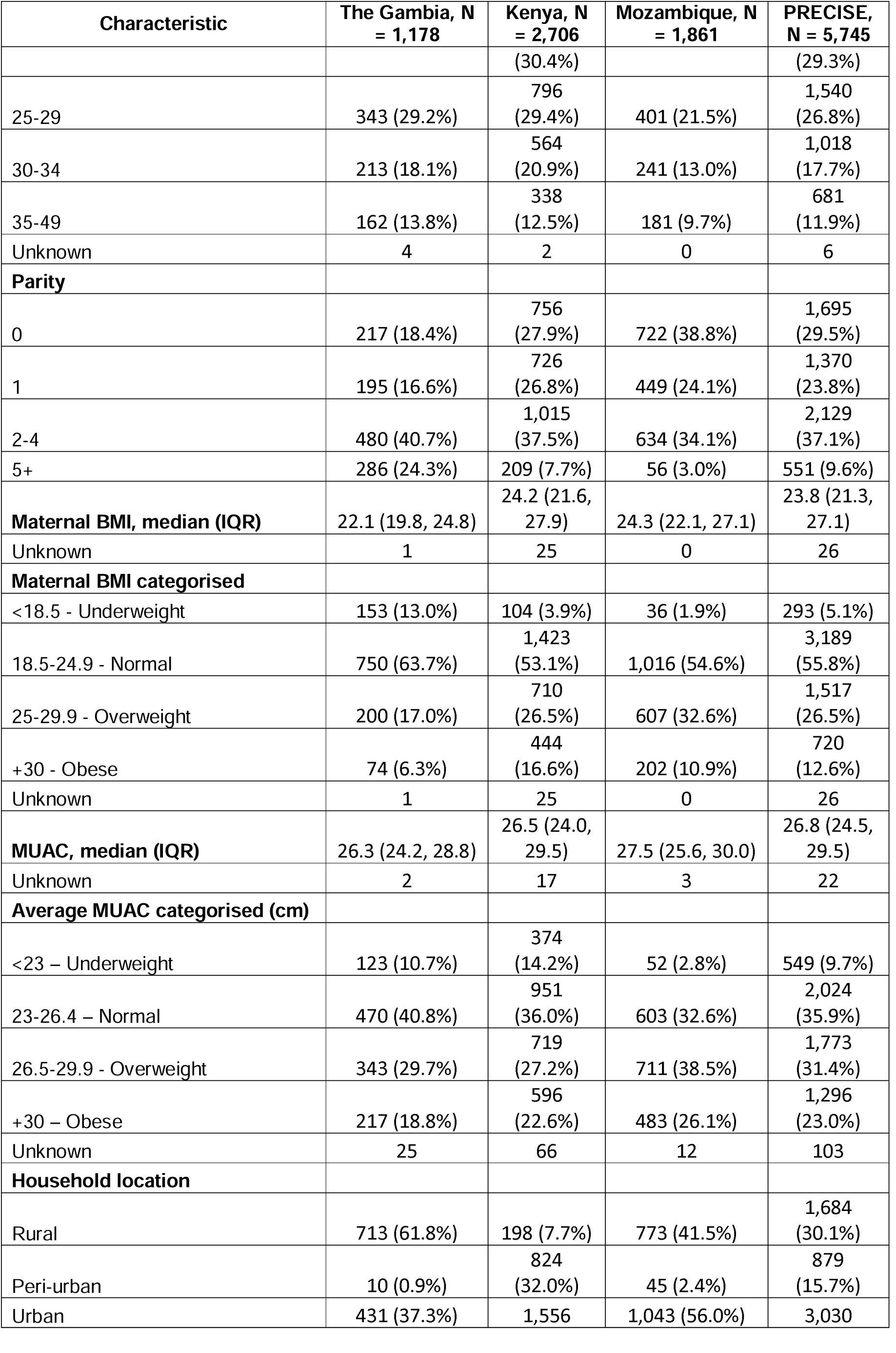
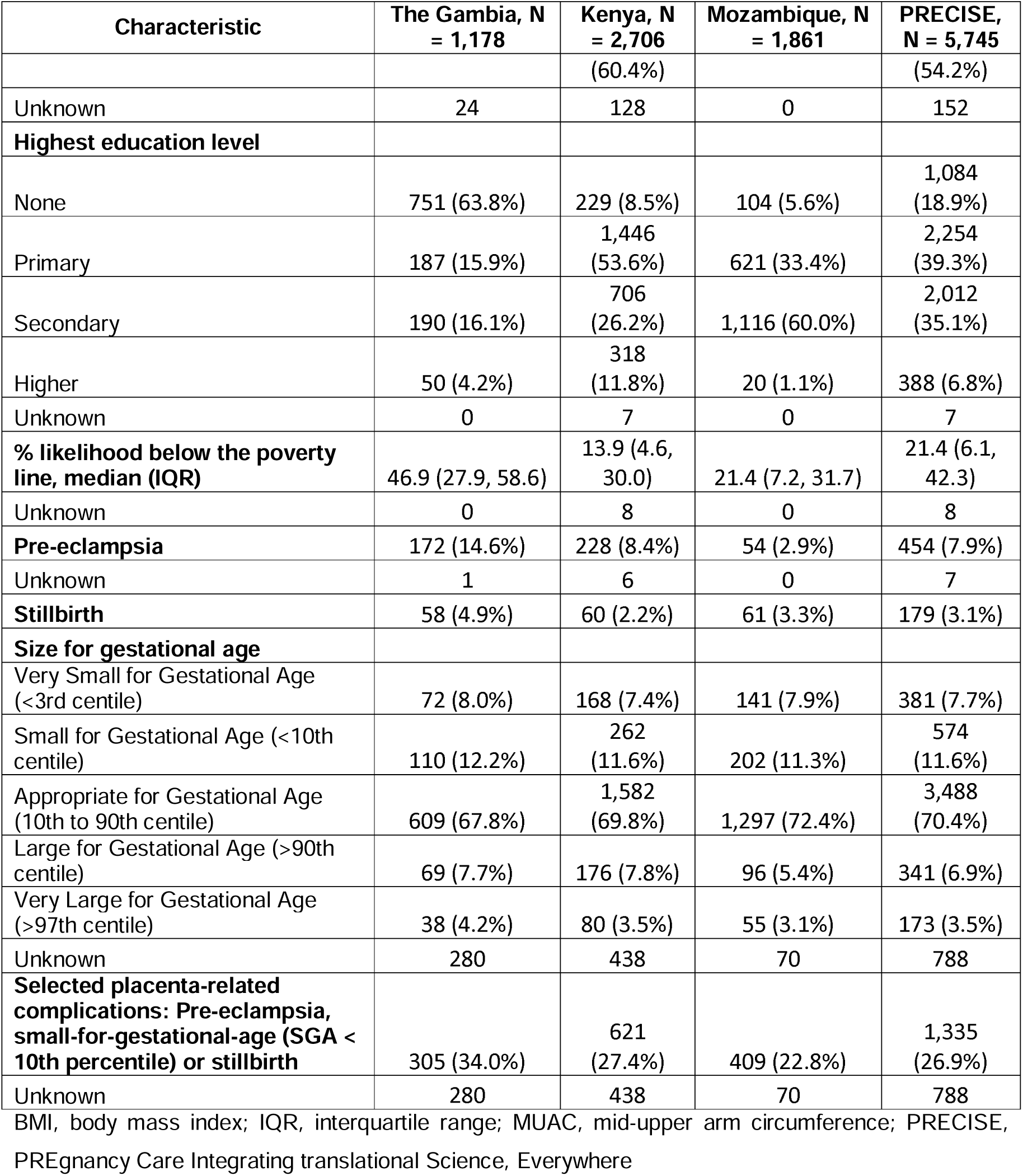
Characteristics of the PRECISE cohort and prevalence of selected placenta-related complications in The Gambia, Kenya and Mozambique.

| Characteristic | The Gambia, N = 1,178 | Kenya, N = 2,706 | Mozambique, N = 1,861 | PRECISE, N = 5,745 |
| --- | --- | --- | --- | --- |
| **Age at enrolment, median (IQR)** | 26.0 (22.0, 31.0) | 26.0 (23.0, 31.0) | 23.0 (19.0, 29.0) | 26.0 (21.0, 31.0) |
| Unknown | 4 | 2 | 0 | 6 |
| **Age at enrolment (years)** |  |  |  |  |
| 15-19 | 145 (12.4%) | 183 (6.8%) | 489 (26.3%) | 817 (14.2%) |
| 20-24 | 311 (26.5%) | 823 | 549 (29.5%) | 1,683 |

| Characteristic | The Gambia, N<br>= 1,178 | Kenya, N<br>= 2,706 | Mozambique, N<br>= 1,861 | PRECISE,<br>N = 5,745 |
| --- | --- | --- | --- | --- |
|  |  | (30.4%) |  | (29.3%) |
| 25-29 | 343 (29.2%) | 796<br>(29.4%) | 401 (21.5%) | 1,540<br>(26.8%) |
| 30-34 | 213 (18.1%) | 564<br>(20.9%) | 241 (13.0%) | 1,018<br>(17.7%) |
| 35-49 | 162 (13.8%) | 338<br>(12.5%) | 181 (9.7%) | 681<br>(11.9%) |
| Unknown | 4 | 2 | 0 | 6 |
| **Parity** |  |  |  |  |
| 0 | 217 (18.4%) | 756<br>(27.9%) | 722 (38.8%) | 1,695<br>(29.5%) |
| 1 | 195 (16.6%) | 726<br>(26.8%) | 449 (24.1%) | 1,370<br>(23.8%) |
| 2-4 | 480 (40.7%) | 1,015<br>(37.5%) | 634 (34.1%) | 2,129<br>(37.1%) |
| 5+ | 286 (24.3%) | 209 (7.7%) | 56 (3.0%) | 551 (9.6%) |
| **Maternal BMI, median (IQR)** | 22.1 (19.8, 24.8) | 24.2 (21.6,<br>27.9) | 24.3 (22.1, 27.1) | 23.8 (21.3,<br>27.1) |
| Unknown | 1 | 25 | 0 | 26 |
| **Maternal BMI categorised** |  |  |  |  |
| <18.5 - Underweight | 153 (13.0%) | 104 (3.9%) | 36 (1.9%) | 293 (5.1%) |
| 18.5-24.9 - Normal | 750 (63.7%) | 1,423<br>(53.1%) | 1,016 (54.6%) | 3,189<br>(55.8%) |
| 25-29.9 - Overweight | 200 (17.0%) | 710<br>(26.5%) | 607 (32.6%) | 1,517<br>(26.5%) |
| +30 - Obese | 74 (6.3%) | 444<br>(16.6%) | 202 (10.9%) | 720<br>(12.6%) |
| Unknown | 1 | 25 | 0 | 26 |
| **MUAC, median (IQR)** | 26.3 (24.2, 28.8) | 26.5 (24.0,<br>29.5) | 27.5 (25.6, 30.0) | 26.8 (24.5,<br>29.5) |
| Unknown | 2 | 17 | 3 | 22 |
| **Average MUAC categorised (cm)** |  |  |  |  |
| <23 – Underweight | 123 (10.7%) | 374<br>(14.2%) | 52 (2.8%) | 549 (9.7%) |
| 23-26.4 – Normal | 470 (40.8%) | 951<br>(36.0%) | 603 (32.6%) | 2,024<br>(35.9%) |
| 26.5-29.9 - Overweight | 343 (29.7%) | 719<br>(27.2%) | 711 (38.5%) | 1,773<br>(31.4%) |
| +30 – Obese | 217 (18.8%) | 596<br>(22.6%) | 483 (26.1%) | 1,296<br>(23.0%) |
| Unknown | 25 | 66 | 12 | 103 |
| **Household location** |  |  |  |  |
| Rural | 713 (61.8%) | 198 (7.7%) | 773 (41.5%) | 1,684<br>(30.1%) |
| Peri-urban | 10 (0.9%) | 824<br>(32.0%) | 45 (2.4%) | 879<br>(15.7%) |
| Urban | 431 (37.3%) | 1,556 | 1,043 (56.0%) | 3,030 |
|  |  | (60.4%) |  | (54.2%) |
| Unknown | 24 | 128 | 0 | 152 |
| **Highest education level** |  |  |  |  |
| None | 751 (63.8%) | 229 (8.5%) | 104 (5.6%) | 1,084<br>(18.9%) |
| Primary | 187 (15.9%) | 1,446<br>(53.6%) | 621 (33.4%) | 2,254<br>(39.3%) |
| Secondary | 190 (16.1%) | 706<br>(26.2%) | 1,116 (60.0%) | 2,012<br>(35.1%) |
| Higher | 50 (4.2%) | 318<br>(11.8%) | 20 (1.1%) | 388 (6.8%) |
| Unknown | 0 | 7 | 0 | 7 |
| **% likelihood below the poverty**<br>**line, median (IQR)** | 46.9 (27.9, 58.6) | 13.9 (4.6,<br>30.0) | 21.4 (7.2, 31.7) | 21.4 (6.1,<br>42.3) |
| Unknown | 0 | 8 | 0 | 8 |
| **Pre-eclampsia** | 172 (14.6%) | 228 (8.4%) | 54 (2.9%) | 454 (7.9%) |
| Unknown | 1 | 6 | 0 | 7 |
| **Stillbirth** | 58 (4.9%) | 60 (2.2%) | 61 (3.3%) | 179 (3.1%) |
| **Size for gestational age** |  |  |  |  |
| Very Small for Gestational Age<br>(<3rd centile) | 72 (8.0%) | 168 (7.4%) | 141 (7.9%) | 381 (7.7%) |
| Small for Gestational Age (<10th<br>centile) | 110 (12.2%) | 262<br>(11.6%) | 202 (11.3%) | 574<br>(11.6%) |
| Appropriate for Gestational Age<br>(10th to 90th centile) | 609 (67.8%) | 1,582<br>(69.8%) | 1,297 (72.4%) | 3,488<br>(70.4%) |
| Large for Gestational Age (>90th<br>centile) | 69 (7.7%) | 176 (7.8%) | 96 (5.4%) | 341 (6.9%) |
| Very Large for Gestational Age<br>(>97th centile) | 38 (4.2%) | 80 (3.5%) | 55 (3.1%) | 173 (3.5%) |
| Unknown | 280 | 438 | 70 | 788 |
| **Selected placenta-related**<br>**complications: Pre-eclampsia,**<br>**small-for-gestational-age (SGA <**<br>**10th percentile) or stillbirth** | 305 (34.0%) | 621<br>(27.4%) | 409 (22.8%) | 1,335<br>(26.9%) |
| Unknown | 280 | 438 | 70 | 788 |

BMI, body mass index; IQR, interquartile range; MUAC, mid-upper arm circumference; PRECISE, PREgnancy Care Integrating translational Science, Everywhere

The Gambia had the highest proportion of women without formal education (64%), compared with Kenya (9%) and Mozambique (6%). In terms of residence, 60% of Kenyan women lived in urban areas and 32% in peri-urban areas. In contrast, 37% of women in The Gambia resided in urban areas and 61% in rural areas. In Mozambique, 56% of women lived in urban areas, while 42% lived in rural areas.

### Prevalence of selected placenta-related complications

Overall, 8% of women had pregnancies complicated by pre-eclampsia, 3% experienced a stillbirth, and 19% had an SGA baby (<10^th^ percentile), with 27% having at least one of these selected placenta-related complications. Overall, Selected placenta-related complications were highest in the Gambia (34%), followed by Kenya (27%) and Mozambique (23%).

### Water, sanitation and hygiene

Across the PRECISE cohort, piped water into the dwelling/plot was the most common source of drinking water (50%). This was highest in Mozambique (71%), compared with Kenya (45%) and The Gambia (29%). Few houses (3%) used an alternative drinking or non-drinking water source. The median time to collect water, if it was collected outside the household, was 5 minutes (IQR: 2–10) (Table 2).

**Table 2.** Water, sanitation and hygiene facilities and practices in the households of pregnant women in the PRECISE cohort in The Gambia, Kenya and Mozambique.

| Variable | The Gambia,<br>N = 1,178 | Kenya,<br>N = 2,706 | Mozambique,<br>N = 1,861 | PRECISE,<br>N = 5,745 |
| --- | --- | --- | --- | --- |
| **Source of drinking water** |  |  |  |  |
| Piped water into dwelling/plot | 341 (29.1%) | 1,204 (44.7%) | 1,320 (71.0%) | 2,865 (50.1%) |
| Other improved water source (piped water into neighbour's dwelling or public tap, boreholes or tubewells, protected dug wells, protected springs, rainwater, and packaged or delivered water) | 792 (67.6%) | 1,396 (51.9%) | 347 (18.7%) | 2,535 (44.3%) |
| Unimproved water source (surface water, unprotected well or spring) | 39 (3.3%) | 91 (3.4%) | 191 (10.3%) | 321 (5.6%) |
| Unknown | 6 | 15 | 3 | 24 |
| **Source of non-drinking water (Used for cooking/washing/personal hygiene)** |  |  |  |  |
| Piped water into dwelling/plot | 349 (29.7%) | 1,228 (45.6%) | 1,309 (70.5%) | 2,886 (50.4%) |
| Other improved water source (piped water into neighbour's dwelling or public tap, boreholes or tubewells, protected dug wells, protected springs, rainwater, | 767 (65.3%) | 1,358 (50.4%) | 354 (19.1%) | 2,479 (43.3%) |

| **Variable** | **The Gambia,**<br>**N = 1,178** | **Kenya,**<br>**N = 2,706** | **Mozambique,**<br>**N = 1,861** | **PRECISE,**<br>**N = 5,745** |
| --- | --- | --- | --- | --- |
| and packaged or delivered water) |  |  |  |  |
| Unimproved water source (surface water, unprotected well or spring) | 58 (4.9%) | 108 (4.0%) | 195 (10.5%) | 361 (6.3%) |
| Unknown | 4 | 12 | 3 | 19 |
| **Alternative drinking and non-drinking water source** | 37 (3.2%) | 70 (2.6%) | 88 (4.7%) | 195 (3.4%) |
| Unknown | 6 | 15 | 4 | 25 |
| **Location of water source** |  |  |  |  |
| In own dwelling | 71 (6.0%) | 474 (17.6%) | 215 (11.6%) | 760 (13.3%) |
| In own yard or plot | 326 (27.7%) | 837 (31.0%) | 1,301 (69.9%) | 2,464 (43.0%) |
| Elsewhere | 781 (66.3%) | 1,386 (51.4%) | 344 (18.5%) | 2,511 (43.8%) |
| Unknown | 0 | 9 | 1 | 10 |
| **Time to collect water if collected outside the household, median (IQR)** | 5.0 (3.0, 10.0) | 5.0 (2.0, 10.0) | 3.0 (2.0, 5.0) | 5.0 (2.0, 10.0) |
| **Sanitation facility** |  |  |  |  |
| Improved (flush toilets connected to piped sewer systems, septic tanks or pit latrines; pit latrines with slabs, composting toilets) | 727 (61.9%) | 2,351 (87.3%) | 1,490 (80.5%) | 4,568 (79.9%) |
| Unimproved (pit latrines without a slab or platform, hanging latrines, bucket latrines or open defecation) | 448 (38.1%) | 342 (12.7%) | 360 (19.5%) | 1,150 (20.1%) |
| Unknown | 3 | 13 | 11 | 27 |
| **Sanitation facility location** |  |  |  |  |
| In own dwelling | 268 (22.8%) | 724 (26.8%) | 194 (10.4%) | 1,186 (20.7%) |
| In own yard or plot | 849 (72.1%) | 1,867 (69.2%) | 1,608 (86.5%) | 4,324 (75.4%) |
| Elsewhere | 61 (5.2%) | 106 (3.9%) | 57 (3.1%) | 224 (3.9%) |
| Unknown | 0 | 9 | 2 | 11 |
| **Share sanitation facility with non-household members** | 421 (35.7%) | 1,484 (55.0%) | 166 (8.9%) | 2,071 (36.1%) |
| Unknown | 0 | 9 | 6 | 15 |
| **Handwashing facility** |  |  |  |  |
| Fixed facility (sink/tap) in dwelling | 20 (1.7%) | 352 (13.2%) | 90 (4.8%) | 462 (8.1%) |
| Fixed facility (sink/tap) in yard/plot | 20 (1.7%) | 187 (7.0%) | 110 (5.9%) | 317 (5.6%) |
| Mobile object (bucket/jug/kettle) | 1,065 (91.0%) | 1,731 (65.0%) | 1,653 (89.0%) | 4,449 (78.2%) |
| No handwashing place in dwelling or yard/plot | 65 (5.6%) | 392 (14.7%) | 4 (0.2%) | 461 (8.1%) |
| Unknown | 8 | 44 | 4 | 56 |

IQR, interquartile range; PRECISE, PREGnancy Care Integrating translational Science, Everywhere

For sanitation, households predominantly had improved sanitation facilities (e.g. flush toilets connected to piped sewer systems, septic tanks or pit latrines; pit latrines with slabs, composting toilets) (80%) compared with unimproved facilities (pit latrines without a slab or platform, hanging latrines, bucket latrines or open defecation) (20%). Use of unimproved facilities was highest in The Gambia (38%) compared with Mozambique (20%) and Kenya (13%). Across all countries, 76% of households had a sanitation facility located in the yard or plot. Overall, 36% of households shared their sanitation facility, but this differed markedly between countries, with 55% in Kenya, 36% in The Gambia and 9% in Mozambique.

Most households in all countries used a mobile object (bucket/jug/kettle) for handwashing (78%). A higher proportion of households in Kenya had no handwashing facility located in the dwelling or yard/plot (15%) compared with The Gambia (6%) and Mozambique (0.2%).

### Joint Monitoring Programme (JMP) ranked categories and national average

In all three countries, the proportion of households with access to at least basic water services exceeded the national average according to the JMP. In The Gambia, 95% of rural households and 96% of urban households in the PRECISE cohort had access to basic water services. In Kenya, the figures were 95% for rural and urban households, while in Mozambique, 83% of rural households and 93% of urban households had access to basic water services (Table S4).

Access to basic sanitation facilities varied across the three countries compared with national averages. In The Gambia, 44% of rural households had access to basic sanitation, compared with the national average (24%) (13). However, in urban households, 44% of households had access to basic sanitation compared with the national average (61%). Rural (35%) and urban (40%) households in Kenya had similar access to basic sanitation facilities compared to the rural (35%) and urban (40%) national average (13). In Mozambique, 74% of rural households had access to basic sanitation, compared with the national average (23%) (13). In urban households, 73% of households had access to basic sanitation compared with the national average (61%) (13) (Table S4). The survey question did not specify whether soap and water were available in the household, so it was not possible to categorise the data into basic hygiene levels.

### Association between WASH and selected placenta-related complications in women

Most WASH variables showed some evidence of an association with grouped selected placenta-related complications (p-value < 0.2), except for alternative water sources for drinking and non-drinking water and for sanitation facility location (Table 3). Drinking water source, non-drinking water source, and source location were correlated (r^2^ > 0.5), with non-drinking water source retained in the analysis due to it having the lowest AIC (Figure S1). Given the high correlation between drinking and non-drinking water sources (Cramer’s V = 0.92), we regard the retained variable as representing both.

**Table 3.**
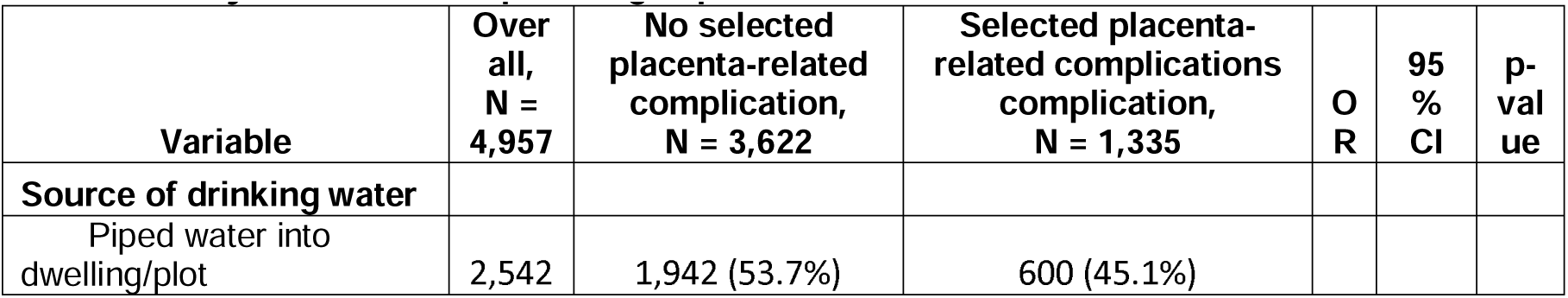
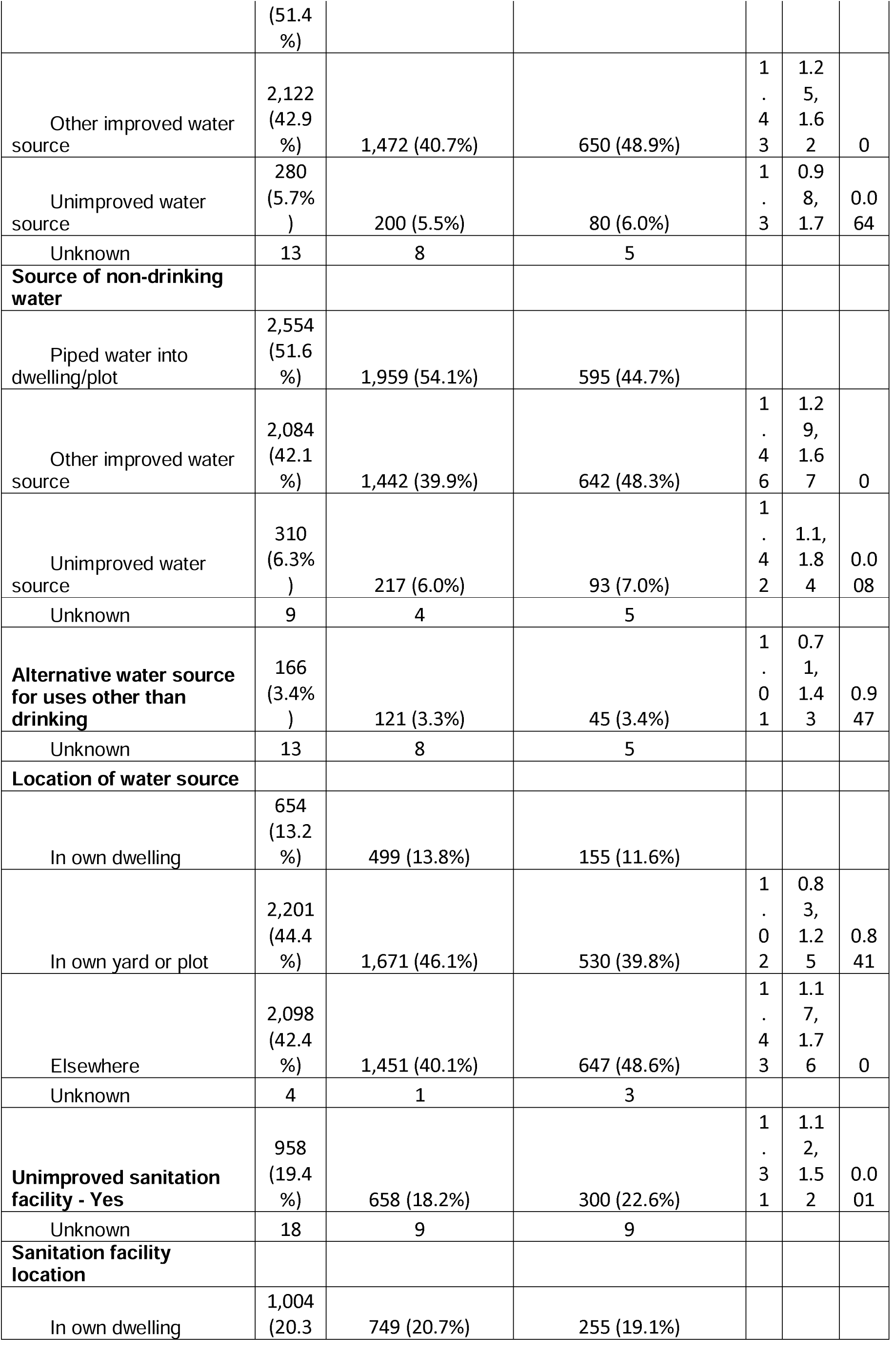
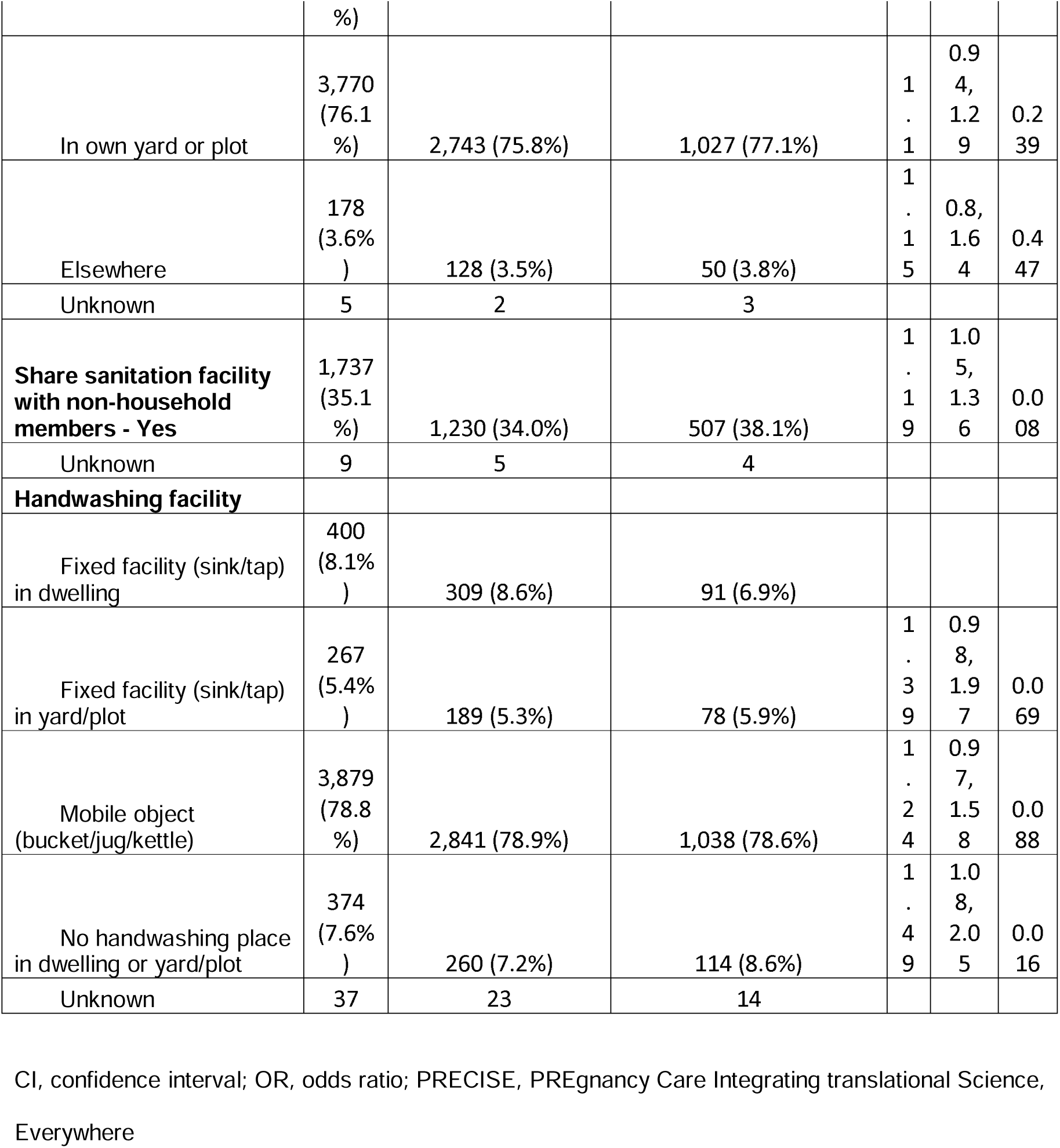
Univariate binomial logistic regression model of 7 WASH variables and their association with selected placenta-related complications in the PRECISE cohort in The Gambia, Kenya and Mozambique using imputed dataset.

Two WASH variables were retained in the multivariate regression (Table 4). After adjusting for a priori confounders and covariates, women relying on other improved water sources had higher odds of experiencing selected placenta-related complications compared to those with piped water in their homes (adjusted odds ratio (aOR) 1.36 [95% confidence interval (CI) 1.18, 1.57], p < 0.001)). The use of unimproved sanitation facilities, compared with improved sanitation facilities, was also retained in the model, but this had a weak association with selected placenta-related complications (1.17 [0.99, 1.39], p = 0.073).

**Table 4.** Multivariate binomial logistic regression model of two WASH variables and their association with selected placenta-related complications in the PRECISE cohort in The Gambia, Kenya and Mozambique.

|  | Unadjusted analyses |  |  | Adjusted analyses^ |  |  |
| --- | --- | --- | --- | --- | --- | --- |
| Variable | OR | 95% CI | p-value | OR | 95% CI | p-value |
| **Source of drinking and cooking water** |  |  |  |  |  |  |
| Piped water into dwelling/plot | — |  |  | — |  |  |
| Other improved water source | 1.43 | 1.25, 1.63 | <0.001 | 1.36 | 1.18, 1.57 | <0.001 |
| Unimproved water source | 1.36 | 1.05, 1.77 | 0.02 | 1.19 | 0.9, 1.57 | 0.219 |
| **Unimproved sanitation facility** | 1.22 | 1.05, 1.43 | 0.011 | 1.17 | 0.99, 1.39 | 0.073 |

OR, odds ratio; CI, confidence interval; PRECISE, PREgnancy Care Integrating translational Science, Everywhere
^Adjusted for: age at enrolment, parity, average MUAC <23 cm (Underweight), household located in rural area, education, % likelihood below the poverty line and season at conception.

### Disaggregated results by country

At the individual country level, compared with piped water into the household, women relying on other improved water sources had higher odds of experiencing selected placenta-related complications in The Gambia (aOR 1.52 [95% CI 1.01, 1.97], p = 0.034) and Kenya (1.29 [1.04, 1.59], p = 0.019), but not in Mozambique. However, unimproved sanitation facilities, compared with improved facilities, were at increased odds of selected placenta-related complications (1.35 [1.02, 1.80], p = 0.038) in Mozambique, but not in The Gambia and Kenya (Table S6).

### Disaggregated results by placenta-related complications

For individual placenta-related complications, compared with women with piped water into the household, other improved water sources were associated with increased odds of pre-eclampsia (aOR 1.42 [95% CI 1.13, 1.77], p = 0.002). Other improved water sources (1.23 [1.05, 1.44], p = 0.001), and unimproved sanitation facilities (1.27 [1.05, 1.53], p = 0.013) were both associated with increased odds of SGA (Table S7). There was no evidence of either the water source or sanitation facility being associated with stillbirth, but this was likely due to a lack of power, with a stillbirth rate of 3% across the entire PRECISE cohort.

## Discussion

This study assessed the WASH status within the homes and communities of pregnant women and the associations with selected placenta-related complications in The Gambia, Kenya and Mozambique. Although studies have explored some of these associations, the effect of WASH on combined placenta-related complications has not been studied. In our study, 27% of women had one or more of the following placenta-related complications: pre-eclampsia, stillbirth or SGA. In both unadjusted and adjusted multivariate models, women with other improved water sources, compared to those with piped water into their dwellings, were associated with increased odds of selected placenta-related complications.

This study found that the odds of selected placenta-related complications increased by 36% for women who had other improved water sources, compared with water piped into their dwellings, after adjusting for confounders. Using an unimproved water source was found to be associated with SGA, but not the grouped pregnancy complication outcome. These findings align with those from other research. In Sierra Leone, using a well or borehole as the main source of drinking water was linked to higher odds of pre-eclampsia/eclampsia (23). However, it is unclear in this study whether this was in comparison with other improved water sources or unimproved water sources. In India, groundwater, compared with piped water, was associated with LBW and very low birth weight (VLBW) (24). Similarly, a pooled analysis in the African Great Lakes region (covering Burundi, the Democratic Republic of Congo, Kenya, Rwanda, Tanzania, and Uganda) found that drinking water from an unimproved source away from the dwelling was associated with stillbirth (6). However, in Nepal, (8) only weak evidence of associations between drinking water sources and stillbirth was found, although it should be noted that Akombi et al. (6) and Ghimire et al. (8) only adjusted for clustering and sampling weight.

Use of unimproved sanitation facilities, including open defecation, compared with improved sanitation facilities, was associated with increased odds of selected placenta-related complications in Mozambique, but not in The Gambia or Kenya. In comparison, a study in India found unimproved sanitation was associated with LBW and VLBW (24). In Nepal, studies have found associations with unimproved sanitation and open defecation with perinatal mortality and stillbirth (8, 25). In rural India, Padhi et al. (9) identified self-reported open defecation to be associated, after adjustment, with increased odds of adverse pregnancy outcomes, defined as an event of preterm birth, LBW, spontaneous abortion or stillbirth. Our study could not explore the associations between open defecation and selected placenta-related complications due to the low number of participants who reported practicing open defecation.

Both water sources outside of the household, including most improved sources and all unimproved sources, and unimproved sanitation facilities pose greater risks to pregnant women (Figure 3). Water sources outside the household have a higher likelihood of contamination, including inorganic contaminants or infectious pathogens that pose risks during pregnancy. Water collected from sources outside the household has to be carried and stored within the household, which can introduce contamination, while unimproved sources do not adequately protect against external contamination. Contaminated water can be ingested by pregnant women, or contaminate food, soil, hands and, fomites, or flies (4) (Figure 3). Unimproved sanitation facilities do not hygienically separate human faeces from contact, resulting in unsafe containment that can lead to the transmission of pathogens from one person to another. Acute maternal gastro-intestinal infections, including *Listeria monocytogenes* and *Escherichia coli*, can be introduced through both these pathways during pregnancy and may contribute to various adverse outcomes, including pre-eclampsia, stillbirth and LBW (5, 26, 27). In China, preterm birth, fetal distress and LBW were significantly higher in pregnant women infected with water-borne? Hepatitis E virus than those who were not (28).

Collecting water from sources located away from the household may pose health risks to pregnant women due to the increased physical burden (Figure 3). Women are often primarily responsible for water collection (29), and given the observed association with water sources located outside the household or premises, it is likely that the study participants had to travel to access water. However, this study did not determine whether the women themselves were collecting the water or how frequently. A systematic review of the effects of occupational activities during pregnancy on outcomes highlights the limited research currently available, particularly concerning lifting weights and placenta-related complications (10). Only one study investigated stillbirth and heavy lifting but found no association (30). In addition, the review found a link between lifting weights (≥11 kg) and increased odds of pre-eclampsia, although there was no association between prolonged standing or walking and pre-eclampsia (10). Meta-analysis indicated no association between lifting weights (≥11 kg) and SGA infants, although prolonged standing and walking were associated with increased odds of SGA, despite low certainty (10). The pathways through which heavy lifting could lead to placenta-related complications are not well understood. Prolonged standing is linked to decreased cardiac output and plasma volume, which may also impact fetal growth (31).

We observed notable disparities in the associations between country-specific factors and selected placenta-related complications. In The Gambia, only access to an improved water source (compared with piped water directly into the household) was significantly associated with selected placenta-related complications. Conversely, in Kenya, both improved and unimproved water sources were linked to higher odds of selected placenta-related complications when compared to household piped water. Interestingly, in both The Gambia and Kenya, no significant associations were found between sanitation facilities and selected placenta-related complications. In Mozambique, on the other hand, unimproved sanitation facilities were associated with increased odds of selected placenta-related complications, whereas water sources showed no significant impact.

These country-specific differences may stem from a variety of factors, including variations in community-level WASH access, affordability, availability, accessibility, water quality, and local behavioural practices related to water usage and sanitation. For example, even “improved” water sources may still be subject to contamination or inconsistent quality (3, 32). Additionally, cultural practices around water storage and sanitation could influence exposure to pathogens, thereby affecting maternal health outcomes differentially across regions.

## Limitations

As this study uses data from a cohort study, we cannot determine the causality between WASH and selected placenta-related complications. The association with other placenta-related complications is also important. Additionally, WASH conditions were only assessed once and at enrolment, so failing to capture any changes in access to WASH facilities throughout pregnancy. Such changes could introduce new risks or mitigate them. The questionnaire was also self-reported which may introduce recall and reporting bias. The study was not designed to be nationally representative of the countries involved, which limits the generalisability of the results.

The questionnaire used also had limitations, offering a restricted set of questions. Further exploration into areas such as water storage, water quantity, menstrual health practices, and handwashing materials could provide better insights into when and how risks are introduced. Additionally, structured observations may offer a more nuanced understanding of WASH-related behaviours and their direct impact on women, beyond the household level. Some variables in the study did not accurately reflect household practices. For instance, questions about water collection did not specify whether the mother or another household member collected the water. Similarly, hygiene questions did not clarify whether soap and water were used for handwashing and if performed correctly.

Furthermore, the questionnaire did not identify the specific WASH-related pathways through which contamination may have reached pregnant women. Environmental samples, including water, food, soil, hand swabs, fomites, or flies could have been tested for faecal indicators, such as *E. coli* and total coliforms, to give a clearer understanding of where infection may be introduced and to help develop targeted interventions. Multi-pathogen PCR analyses or other techniques could have been employed to identify individual pathogens, enhancing our understanding of contamination pathways and infection risks for pregnant women.

## Conclusions

A quarter of women enrolled in this study had one or more of the following selected placenta-related complications: pre-eclampsia, stillbirth, or an SGA infant. After adjusting for potential confounders, women using improved drinking water sources other than household piped water, as well as those with unimproved sanitation facilities compared to improved ones, had a higher risk of developing selected placenta-related complications. Overall, our findings are consistent with other studies exploring similar associations, reinforcing our understanding of the interaction of WASH and selected placenta-related complications.

The findings underscore the importance of integrating WASH interventions into health improvement programs globally. Improved access to clean water and sanitation could potentially reduce the incidence of placenta-related complications, thereby improving birth outcomes. WASH interventions could also be used to understand the causality between WASH and placenta-related complications. However, these interventions need to be developed with local context and practices in mind as WASH behaviours often differ greatly between populations. Control measures, such as improving water quality through chlorination or advising pregnant women against heavy lifting of water containers, may also help reduce the incidence of placenta-related complications. Further robust research is needed to confirm the associations identified in this study and to co-design context-appropriate improvements to all aspects of WASH services in collaboration with communities.

## Data Availability

The data that support the findings of this study are available from the Research Programme Manager MLV, on reasonable request. The guidelines for data and sample access requests for the PRECISE network can be found here: https://precisenetwork.org/precise/data-and-sample-access/

## References

1. von Dadelszen P, Magee LA. Preventing deaths due to the hypertensive disorders of pregnancy. Best Practice & Research Clinical Obstetrics & Gynaecology. 2016;36:83–102.

2. Ross I, Bick S, Ayieko P, Dreibelbis R, Wolf J, Freeman MC, et al. Effectiveness of handwashing with soap for preventing acute respiratory infections in low-income and middle-income countries: a systematic review and meta-analysis. The Lancet. 2023;401(10389):1681–90.

3. Wolf J, Hubbard S, Brauer M, Ambelu A, Arnold BF, Bain R, et al. Effectiveness of interventions to improve drinking water, sanitation, and handwashing with soap on risk of diarrhoeal disease in children in low-income and middle-income settings: a systematic review and meta-analysis. The Lancet. 2022;400(10345):48–59.

4. Campbell OMR, Benova L, Gon G, Afsana K, Cumming O. Getting the basic rights – the role of water, sanitation and hygiene in maternal and reproductive health: a conceptual framework. Tropical Medicine & International Health. 2015;20(3):252–67.

5. Kumar M, Saadaoui M, Al Khodor S. Infections and Pregnancy: Effects on Maternal and Child Health. Frontiers in Cellular and Infection Microbiology. 2022;12.

6. Akombi BJ, Ghimire PR, Agho KE, Renzaho AM. Stillbirth in the African Great Lakes region: A pooled analysis of Demographic and Health Surveys. PLoS One. 2018;13(8):e0202603.

7. Kayode GA, Amoakoh-Coleman M, Agyepong IA, Ansah E, Grobbee DE, Klipstein-Grobusch K. Contextual risk factors for low birth weight: a multilevel analysis. PLoS One. 2014;9(10):e109333.

8. Ghimire PR, Agho KE, Renzaho AMN, Nisha MK, Dibley M, Raynes-Greenow C. Factors associated with perinatal mortality in Nepal: evidence from Nepal demographic and health survey 2001-2016. BMC Pregnancy Childbirth. 2019;19(1):88.

9. Padhi BK, Baker KK, Dutta A, Cumming O, Freeman MC, Satpathy R, et al. Risk of Adverse Pregnancy Outcomes among Women Practicing Poor Sanitation in Rural India: A Population-Based Prospective Cohort Study. PLOS Medicine. 2015;12(7):e1001851.

10. Cai C, Vandermeer B, Khurana R, Nerenberg K, Featherstone R, Sebastianski M, et al. The impact of occupational activities during pregnancy on pregnancy outcomes: a systematic review and metaanalysis. American Journal of Obstetrics and Gynecology. 2020;222(3):224–38.

11. Meierhofer R, Tomberge VMJ, Inauen J, Shrestha A. Water carrying in hills of Nepal–associations with women’s musculoskeletal disorders, uterine prolapse, and spontaneous abortions. PLOS ONE. 2022;17(6):e0269926.

12. Craik R, Akuze J, Volvert M-L, Blencowe H, Mukhanya M, Makanga PT, et al. PREgnancy Care Integrating translational Science, Everywhere (PRECISE): a prospective cohort study of African pregnant and non-pregnant women to investigate placental disorders – cohort profile. BMJ Open. 2025;15(5):e091831.

13. World Health Organization, United Nations Children’s Fund. The WHO/UNICEF joint monitoring programme estimates on WASH. 2023 [Available from: https://washdata.org/data.

14. von Dadelszen P, Flint-O’Kane M, Poston L, Craik R, Russell D, Tribe RM, et al. The PRECISE (PREgnancy Care Integrating translational Science, Everywhere) Network’s first protocol: deep phenotyping in three sub-Saharan African countries. Reproductive Health. 2020;17(S1).

15. Magee LA, Brown MA, Hall DR, Gupte S, Hennessy A, Karumanchi SA, et al. The 2021 International Society for the Study of Hypertension in Pregnancy classification, diagnosis & management recommendations for international practice. Pregnancy Hypertension. 2022;27:148–69.

16. Villar J, Ismail LC, Victora CG, Ohuma EO, Bertino E, Altman DG, et al. International standards for newborn weight, length, and head circumference by gestational age and sex: the Newborn Cross-Sectional Study of the INTERGROWTH-21st Project. The Lancet. 2014;384(9946):857–68.

17. WHO Expert Committee on Health Statistics. Expert Committee on Health Statistics: report on the second session, Geneva, 18-21 April 1950, including reports on the first sessions of the Subcommittees on Definition of Stillbirth, Registration of Cases of Cancer, Hospital Statistics. Geneva: World Health Organization; 1950.

18. Buuren SV, Groothuis-Oudshoorn K. mice: Multivariate Imputation by Chained Equations in R. Journal of Statistical Software. 2011;45(3).

19. Posit team. RStudio: Integrated development environment for R. Boston, MA: Posit Software, PBC; 2024.

20. Venables WN, Ripley BD. Modern Applied Statistics with S. Fourth ed. New York: Springer; 2002.

21. Bursac Z, Gauss CH, Williams DK, Hosmer DW. Purposeful selection of variables in logistic regression. Source Code for Biology and Medicine. 2008;3(1):17.

22. Fox J, Weisberg S. An R companion to applied regression. Third ed. Thousand Oaks, CA: Sage; 2018.

23. Stitterich N, Shepherd J, Koroma MM, Theuring S. Risk factors for preeclampsia and eclampsia at a main referral maternity hospital in Freetown, Sierra Leone: a case-control study. BMC Pregnancy and Childbirth. 2021;21(1).

24. Biswas S, Mondal S, Banerjee A, Alam A, Satpati L. Investigating the association between floods and low birth weight in India: Using the geospatial approach. Science of The Total Environment. 2024;912:169593.

25. Ghimire PR, Agho KE, Renzaho A, Christou A, Nisha MK, Dibley M, et al. Socio-economic predictors of stillbirths in Nepal (2001-2011). PLOS ONE. 2017;12(7):e0181332.

26. Xie F, Turvey SE, Williams MA, Mor G, Von Dadelszen P. Toll-Like Receptor Signaling and Pre-Eclampsia. American Journal of Reproductive Immunology. 2010;63(1):7–16.

27. von Dadelszen P, Magee LA. Could an infectious trigger explain the differential maternal response to the shared placental pathology of preeclampsia and normotensive intrauterine growth restriction? Acta Obstetricia et Gynecologica Scandinavica. 2002;81(7):642–8.

28. Wen G-P, Wang M-M, Tang Z-M, Liu C, Yu Z-H, Wang Z, et al. Prevalence of Hepatitis E Virus and Its Associated Outcomes among Pregnant Women in China. Pathogens. 2023;12(9):1072.

29. Graham JP, Hirai M, Kim S-S. An Analysis of Water Collection Labor among Women and Children in 24 Sub-Saharan African Countries. PLOS ONE. 2016;11(6):e0155981.

30. Juhl M, Strandberg-Larsen K, Larsen PS, Andersen PK, Svendsen SW, Bonde JP, et al. Occupational lifting during pregnancy and risk of fetal death in a large national cohort study. Scandinavian Journal of Work, Environment & Health. 2013;39(4):335–42.

31. Redman CW. Maternal plasma volume and disorders of pregnancy. BMJ. 1984;288(6422):955-6.

32. Bain R, Cronk R, Wright J, Yang H, Slaymaker T, Bartram J. Fecal Contamination of Drinking-Water in Low– and Middle-Income Countries: A Systematic Review and Meta-Analysis. PLoS Medicine. 2014;11(5):e1001644.

