## Supplementary Information for "Evaluating the association between household water, sanitation and hygiene (WASH) and selected placenta-related complications in The Gambia, Kenya and Mozambique"

**Table S1: The PRECISE Network**

| **In-country teams** | **Members** |
| --- | --- |
| THE GAMBIA: Medical Research Council Unit The Gambia at the London School of Hygiene and Tropical Medicine, Fajara | Umberto D’Alessandro, Anna Roca, Hawanatu Jah, Andrew Prentice, Melisa Martinez-Alvarez, Brahima Diallo, Abdul Sesay, Sambou Suso, Yahaya Idris, Baboucarr Njie, Fatima Touray, Fatoumata Kongira, Modou F.S. Ndure, Gibril Gabbidon, Lawrence Gibba, Abdoulie Bah and Yorro Bah. |
| KENYA: Aga Khan University, Nairobi | Marleen Temmerman, Angela Koech, Patricia Okiro, Geoffrey Omuse, Grace Mwashigadi, Consolata Juma, Joseph Mutunga, Moses Mukhanya, Onesmus Wanje, Isaac Mwaniki, Marvin Ochieng, Emily Mwadime |
| MOZAMBIQUE : Centro de Investigação em Saúde de Manhiça, Manhiça | Esperança Sevene, Corssino Tchavana, Salesio Macuacua, Anifa Vala, Helena Boene, Lazaro Quimice, Sonia Maculuve, Inacio Mandomando |
| **Central co-ordinating team** |  |
| Department of Women and Children’s Health, School of Life Course Sciences, Faculty of Life Sciences and Medicine, King’s College London | Peter von Dadelszen, Laura A. Magee, Rachel Craik, Marie-Laure Volvert, Hiten Mistry, Meriel Flint-O’Kane, Amber Strang, Marina Daniele, Thomas Mendy |
| Donna Russell Consulting | Donna Russell |
| **Co-Investigator team** |  |
| Midlands State University, Zimbabwe | Prestige Tatenda Makanga, Liberty Makacha and Reason Mlambo |
| Kings College London | Lucilla Poston, Rachel Tribe, Sophie Moore, Tatiana Salisbury |
| University of Oxford | Aris Papageorghiou, Alison Noble, Rachel Craik |
| London School of Hygiene and Tropical Medicine | Hannah Blencowe, Veronique Filippi, Joy Lawn, Matt Silver, Joseph Akuze and Ursula Gazeley |
| St George’s, University of London | Judith Cartwright, Guy Whitley, Sanjeev Krishna |
| University of British Columbia | Marianne Vidler, Jing (Larry) Li, Jeff Bone, Mai-Lei (Maggie) Woo Kinshella, Domena Tu, Ash Sandhu, Kelly Pickerill |
| Eduardo Mondlane University, Maputo | Carla Carrilho |
| Imperial College London | Benjamin Barratt |

PRECISE, PREgnancy Care Integrating translational Science, Everywhere

**Table S2: The PRECISE-DYAD Network**

| **In-country teams** | **Members** |
| --- | --- |
| THE GAMBIA: Medical Research Council Unit The Gambia at the London School of Hygiene and Tropical Medicine, Fajara | Umberto D’Alessandro, Anna Roca, Hawanatu Jah, Andrew Prentice, Melisa Martinez-Alvarez, Brahima Diallo, Abdul Sesay, Sambou Suso, Yahaya Idris, Baboucarr Njie, Fatima Touray, Fatoumata Kongira, Modou F.S. Ndure, Gibril Gabbidon, Lawrence Gibba, Abdoulie Bah and Yorro Bah. |
| KENYA: Aga Khan University, Nairobi | Marleen Temmerman, Angela Koech, Patricia Okiro, Geoffrey Omuse, Amina Abubakar, Grace Mwashigadi, Isaac Mwaniki, Joseph Mutunga, Moses Mukhanya, Onesmus Wanje, Marvin Ochieng, and Agnes Mutua, Emily Mwadime |
| MOZAMBIQUE : Centro de Investigação em Saúde de Manhiça, Manhiça | Esperança Sevene, Corssino Tchavana, Salesio Macuacua, Jovito Nunes, Valdemiro Escola and Charfudin Sacoor |
| Central co-ordinating team |  |
| Department of Women and Children’s Health, School of Life Course Sciences, Faculty of Life Sciences and Medicine, King’s College London | Peter von Dadelszen, Laura A. Magee, Rachel Craik, Marie-Laure Volvert, Hiten Mistry, Cristina Escalona, Anne Rerimoi, Giulia Ghillia, Thomas Mendy, |
| **Co-Investigator team** |  |
| Midlands State University, Zimbabwe | Prestige Tatenda Makanga, Liberty Makacha, Reason Mlambo |
| Kings College London | Lucilla Poston, Rachel Tribe, Sophie Moore, Tatiana Taylor Salisbury |
| London School of Hygiene and Tropical Medicine | Hannah Blencowe, Veronique Filippi, Joy LawnJaya Chandna, Joseph Akuze |
| St George’s, University of London | Asma Khalil |
| University of British Columbia | Marianne Vidler, Jing (Larry) Li, Jeff Bone, Mai-Lei (Maggie) Woo Kinshella, Domena Tu, Akshdeep Sandhu, Kelly Pickerill |
| Imperial College London | Benjamin Barratt |
| University of Liverpool/Liverpool School of Tropical Medicine | Melissa Gladstone, Dorcas Magai |
| ISGlobal | Cathryn Tonne, Ariadna Curto, Ariadna Moreno |
| C-Squared Development | Chris Clarke |

PRECISE, PREgnancy Care Integrating translational Science, Everywhere

**Table S3.** Baseline characteristics of participants with missing size-for-gestational age data compared with those that do not in the PRECISE cohort in The Gambia, Kenya and Mozambique.

| **Characteristic** | **SGA** | |
| --- | --- | --- |
|  | **Not missing,**  **N = 4,957 (86.3%)** | **Missing,**  **N = 788 (13.7%)** |
| **Age at enrolment, median (IQR)** | 25 (21, 30) | 26 (22, 31) |
| Unknown | 1 | 5 |
| **Age at enrolment (years)** |  |  |
| 15-19 | 747 (15.1%) | 70 (8.9%) |
| 20-24 | 1,456 (29.4%) | 227 (29.0%) |
| 25-29 | 1,313 (26.5%) | 227 (29.0%) |
| 30-34 | 868 (17.5%) | 150 (19.2%) |
| 35+ | 572 (11.5%) | 109 (13.9%) |
| Unknown | 1 | 5 |
| **Parity** |  |  |
| 0 | 1,510 (30.5%) | 185 (23.5%) |
| 1 | 1,196 (24.1%) | 174 (22.1%) |
| 2-4 | 1,817 (36.7%) | 312 (39.6%) |
| 5+ | 434 (8.8%) | 117 (14.8%) |
| **Maternal BMI, median (IQR)** | 23.9 (21.4, 27.2) | 23.3 (20.7, 26.6) |
| Unknown | 22 | 4 |
| **Maternal BMI categorised** |  |  |
| <18.5 - Underweight | 239 (4.8%) | 54 (6.9%) |
| 18.5-24.9 - Normal | 2,726 (55.2%) | 463 (59.1%) |
| 25-29.9 - Overweight | 1,345 (27.3%) | 172 (21.9%) |
| +30 - Obese | 625 (12.7%) | 95 (12.1%) |
| Unknown | 22 | 4 |
| **Average MUAC, median (IQR)** | 26.8 (24.6, 29.5) | 27.5 (24.3, 29.4) |
| Unknown | 17 | 5 |
| **Average MUAC categorised (cm)** |  |  |
| <23 – Underweight | 469 (9.6%) | 80 (10.4%) |
| 23-26.4 – Normal | 1,733 (35.6%) | 291 (37.8%) |
| 26.5-29.9 - Overweight | 1,538 (31.6%) | 235 (30.5%) |
| +30 – Obese | 1,132 (23.2%) | 164 (21.3%) |
| Unknown | 85 | 18 |
| **Household location** |  |  |
| Rural | 1,447 (29.9%) | 237 (31.5%) |
| Peri-urban | 729 (15.1%) | 150 (19.9%) |
| Urban | 2,665 (55.1%) | 365 (48.5%) |
| Unknown | 116 | 36 |
| **Highest education level** |  |  |
| None | 857 (17.3%) | 227 (29.0%) |
| Primary | 1,920 (38.7%) | 334 (42.7%) |
| Secondary | 1,841 (37.2%) | 171 (21.8%) |
| Higher | 337 (6.8%) | 51 (6.5%) |
| Unknown | 2 | 5 |
| **% likelihood below the poverty line, median (IQR)** | 21.4 (6.1, 37.4) | 27.9 (6.1, 50.8) |
| Unknown | 2 | 6 |
| **Pre-eclampsia** | 410 (8.3%) | 44 (5.6%) |
| Unknown | 3 | 4 |
| **Stillbirth** | 112 (2.3%) | 67 (8.5%) |

BMI, body mass index; IQR, interquartile range; MUAC, mid-upper arm circumference; SGA, small-for-gestational-age

**Table S4.** Comparison of Joint Monitoring Programme 2022 national WASH data and the PRECISE cohort in The Gambia, Kenya and Mozambique.

| **JMP Ladders** | **The Gambia** | | | | **Kenya** | | | | **Mozambique** | | | |
| --- | --- | --- | --- | --- | --- | --- | --- | --- | --- | --- | --- | --- |
|  | **Rural** | | **Urban** | | **Rural** | | **Urban** | | **Rural** | | **Urban** | |
|  | **JMP data, %** | **PRECISE* (N=713)** | **JMP data, %** | **PRECISE (N=431)** | **JMP data, %** | **PRECISE (N = 198)** | **JMP data, %** | **PRECISE (N = 1,556)** | **JMP data,**  **%** | **PRECISE (N = 773)** | **JMP data, %** | **PRECISE**  **(N = 1,043)** |
| **Water** |  |  |  |  |  |  |  |  |  |  |  |  |
| At least basic | 76.4 | 611 (95.0%) | 90.9 | 401 (96.6%) | 53.3 | 186 (94.9%) | 86.4 | 1,448 (94.5%) | 48.3 | 642 (83.4%) | 87.3 | 963 (93.3%) |
| Limited | 12.5 | 2 (0.3%) | 2.2 | 5 (1.2%) | 10.7 | 7 (3.6%) | 3.6 | 61 (4.0%) | 12.5 | 5 (0.6%) | 5.4 | 5 (0.5%) |
| Unimproved | 11.0 | 30 (4.7%) | 6.9 | 9 (2.2%) | 11.5 | 2 (1.0%) | 4.0 | 10 (0.7%) | 23.7 | 122 (15.8%) | 5.3 | 62 (6.0%) |
| Surface water | 0 | 0 (0.0%) | 0.1 | 0 (0.0%) | 24.5 | 1 (0.5%) | 6.0 | 13 (0.8%) | 15.4 | 1 (0.1%) | 2.1 | 2 (0.2%) |
| Unknown |  | 70 |  | 16 |  | 2 |  | 24 |  | 3 |  | 11 |
| **Sanitation** |  |  |  |  |  |  |  |  |  |  |  |  |
| At least basic | 24.0 | 313 (44.0%) | 61.1 | 190 (44.1%) | 35.2 | 70 (35.4%) | 39.8 | 621 (39.9%) | 22.6 | 567 (73.5%) | 61.3 | 753 (73.0%) |
| Limited | 7.9 | 78 (11.0%) | 13.8 | 124 (28.8%) | 15.9 | 109 (55.1%) | 44.9 | 772 (49.6%) | 1.7 | 35 (4.5%) | 10.5 | 97 (9.4%) |
| Unimproved | 68 | 314 (44.1%) | 25 | 115 (26.7%) | 40.2 | 16 (8.1%) | 14.4 | 151 (9.7%) | 47.3 | 168 (21.8%) | 22.9 | 178 (17.3%) |
| OD | 0.0 | 7 (1.0%) | 0.1 | 2 (0.5%) | 8.7 | 3 (1.5%) | 0.9 | 12 (0.8%) | 28.5 | 1 (0.1%) | 5.2 | 3 (0.3%) |
| Unknown |  | 1 |  | 0 |  | 0 |  | 0 |  | 2 |  | 12 |
| **Hygiene** |  |  |  |  |  |  |  |  |  |  |  |  |
| Basic | 11.9 | NA | 13.5 | NA | 34.6 | NA | 44.8 | NA | NA | NA | NA | NA |
| Limited^1^ | 80.2 | 660 (93.4%) | 77.4 | 413 (96.3%) | 27.4 | 158 (80.6%) | 31.3 | 1,329 (86.6%) | NA | 770 (99.9%) | NA | 1,038 (99.7%) |
| No facility | 7.9 | 47 (6.6%) | 9.1 | 16 (3.7%) | 38 | 38 (19.4%) | 23.9 | 206 (13.4%) | NA | 1 (0.1%) | NA | 3 (0.3%) |
| Unknown |  | 6 |  | 2 |  | 2 |  | 21 |  | 2 |  | 2 |

Note: this does not include peri-urban data. All national WASH data sourced from the Joint Monitoring Programme in 2022 (13).

Abbreviations: JMP, Joint Monitoring Programme; OD, open defecation; PRECISE, PREgnancy Care Integrating translational Science, Everywhere

^1^ Survey question did not specify whether soap and water were available in the household, so it was not possible to categorise PRECISE countries into the basic hygiene levels.

**Table S5.** Univariate binomial logistic regression model of 7 WASH variables and their association with selected placenta-related complications in the PRECISE cohort in The Gambia, Kenya and Mozambique with imputed and non-imputed datasets.

|  |  |  |  | **Imputed dataset** | | | **Non-imputed dataset** | | |
| --- | --- | --- | --- | --- | --- | --- | --- | --- | --- |
| **Variable** | **Overall,**  **N = 4,957** | **No selected placenta-related complication,**  **N = 3,622** | **Selected placenta-related complication, N = 1,335** | **OR** | **95% CI** | **p-value** | **OR** | **95% CI** | **p-value** |
| **Source of drinking water** |  |  |  |  |  |  |  |  |  |
| Piped water into dwelling/plot | 2,542 (51.4%) | 1,942 (53.7%) | 600 (45.1%) |  |  |  |  |  |  |
| Other improved water source | 2,122 (42.9%) | 1,472 (40.7%) | 650 (48.9%) | 1.43 | 1.25, 1.62 | <0.001 | 1.43 | 1.26, 1.63 | <0.001 |
| Unimproved water source | 280 (5.7%) | 200 (5.5%) | 80 (6.0%) | 1.3 | 0.98, 1.7 | 0.064 | 1.29 | 0.98, 1.70 | 0.066 |
| Unknown | 13 | 8 | 5 |  |  |  |  |  |  |
| **Source of non-drinking water** |  |  |  |  |  |  |  |  |  |
| Piped water into dwelling/plot | 2,554 (51.6%) | 1,959 (54.1%) | 595 (44.7%) |  |  |  |  |  |  |
| Other improved water source | 2,084 (42.1%) | 1,442 (39.9%) | 642 (48.3%) | 1.46 | 1.29, 1.67 | <0.001 | 1.47 | 1.29, 1.67 | <0.001 |
| Unimproved water source | 310 (6.3%) | 217 (6.0%) | 93 (7.0%) | 1.42 | 1.1, 1.84 | 0.008 | 1.41 | 1.08, 1.82 | 0.009 |
| Unknown | 9 | 4 | 5 |  |  |  |  |  |  |
| **Alternative water source for uses other than drinking** | 166 (3.4%) | 121 (3.3%) | 45 (3.4%) | 1.01 | 0.71, 1.43 | 0.947 | 1.01 | 0.71, 1.42 | 0.951 |
| Unknown | 13 | 8 | 5 |  |  |  |  |  |  |
| **Location of water source** |  |  |  |  |  |  |  |  |  |
| In own dwelling | 654 (13.2%) | 499 (13.8%) | 155 (11.6%) |  |  |  |  |  |  |
| In own yard or plot | 2,201 (44.4%) | 1,671 (46.1%) | 530 (39.8%) | 1.02 | 0.83, 1.25 | 0.841 | 1.02 | 0.83, 1.26 | 0.842 |
| Elsewhere | 2,098 (42.4%) | 1,451 (40.1%) | 647 (48.6%) | 1.43 | 1.17, 1.76 | <0.001 | 1.44 | 1.17, 1.76 | <0.001 |
| Unknown | 4 | 1 | 3 |  |  |  |  |  |  |
| **Unimproved sanitation facility - Yes** | 958 (19.4%) | 658 (18.2%) | 300 (22.6%) | 1.31 | 1.12, 1.52 | 0.001 | 1.31 | 1.12, 1.53 | <0.001 |
| Unknown | 18 | 9 | 9 |  |  |  |  |  |  |
| **Sanitation facility location** |  |  |  |  |  |  |  |  |  |
| In own dwelling | 1,004 (20.3%) | 749 (20.7%) | 255 (19.1%) |  |  |  |  |  |  |
| In own yard or plot | 3,770 (76.1%) | 2,743 (75.8%) | 1,027 (77.1%) | 1.1 | 0.94, 1.29 | 0.239 | 1.10 | 0.94, 1.29 | 0.242 |
| Elsewhere | 178 (3.6%) | 128 (3.5%) | 50 (3.8%) | 1.15 | 0.8, 1.64 | 0.447 | 1.15 | 0.80, 1.63 | 0.450 |
| Unknown | 5 | 2 | 3 |  |  |  |  |  |  |
| **Share sanitation facility with non-household members - Yes** | 1,737 (35.1%) | 1,230 (34.0%) | 507 (38.1%) | 1.19 | 1.05, 1.36 | 0.008 | 1.19 | 1.05, 1.36 | 0.008 |
| Unknown | 9 | 5 | 4 |  |  |  |  |  |  |
| **Handwashing facility** |  |  |  |  |  |  |  |  |  |
| Fixed facility (sink/tap) in dwelling | 400 (8.1%) | 309 (8.6%) | 91 (6.9%) |  |  |  |  |  |  |
| Fixed facility (sink/tap) in yard/plot | 267 (5.4%) | 189 (5.3%) | 78 (5.9%) | 1.39 | 0.98, 1.97 | 0.069 | 1.40 | 0.98, 1.99 | 0.061 |
| Mobile object (bucket/jug/kettle) | 3,879 (78.8%) | 2,841 (78.9%) | 1,038 (78.6%) | 1.24 | 0.97, 1.58 | 0.088 | 1.24 | 0.98, 1.59 | 0.084 |
| No handwashing place in dwelling or yard/plot | 374 (7.6%) | 260 (7.2%) | 114 (8.6%) | 1.49 | 1.08, 2.05 | 0.016 | 1.49 | 1.08, 2.06 | 0.015 |
| Unknown | 37 | 23 | 14 |  |  |  |  |  |  |

Abbreviations: CI, confidence interval; OR, odds ratio; PRECISE, PREgnancy Care Integrating translational Science, Everywhere

**Table S6**. Comparison of the adjusted final multivariate model for the PRECISE cohort by country (The Gambia, Kenya and Mozambique)

|  | **The Gambia (N = 898)** | | | **Kenya (N = 2,268)** | | | **Mozambique (N = 1,791)** | | |
| --- | --- | --- | --- | --- | --- | --- | --- | --- | --- |
| **Variable** | **aOR** | **95% CI** | **p-value** | **aOR** | **95% CI** | **p-value** | **aOR** | **95% CI** | **p-value** |
| **Source of drinking water** |  |  |  |  |  |  |  |  |  |
| Piped water into dwelling/plot | — |  |  | **—** |  |  | — |  |  |
| Other improved water source | 1.52 | 1.03, 2.24 | 0.034 | 1.29 | 1.04, 1.59 | 0.019 | 1.22 | 0.92, 1.62 | 0.168 |
| Unimproved water source | 1.08 | 0.49, 2.38 | 0.854 | 1.36 | 0.79, 2.35 | 0.264 | 1.25 | 0.87, 1.81 | 0.234 |
| **Unimproved sanitation facility** | 1 | 0.73, 1.35 | 0.976 | 1.08 | 0.79, 1.49 | 0.616 | 1.35 | 1.02, 1.8 | 0.038 |

Note: adjusted for: age at enrolment, parity, average MUAC <23 cm (Underweight), household located in rural area, education, % likelihood below the poverty line and season at conception.

Abbreviations: CI, confidence interval; aOR, adjusted odds ratio; PRECISE, PREgnancy Care Integrating translational Science, Everywhere

**Table S7.** Comparison of the adjusted final multivariate model for the PRECISE cohort by pregnancy complication

|  | **Pre-eclampsia (N = 5,738)** | | | **Stillbirth (N = 5,745)** | | | **SGA (N = 4,957)** | | |
| --- | --- | --- | --- | --- | --- | --- | --- | --- | --- |
| **Variable** | **aOR** | **95% CI** | **p-value** | **aOR** | **95% CI** | **p-value** | **aOR** | **95% CI** | **p-value** |
| **Source of drinking water** |  |  |  |  |  |  |  |  |  |
| Piped water into dwelling/plot | — |  |  | **—** |  |  | — |  |  |
| Other improved water source | 1.42 | 1.13, 1.77 | 0.002 | 1.15 | 0.81, 1.63 | 0.426 | 1.23 | 1.05, 1.44 | 0.010 |
| Unimproved water source | 1.12 | 0.73, 1.75 | 0.599 | 1.32 | 0.74, 2.35 | 0.339 | 1.17 | 0.86, 1.58 | 0.312 |
| **Unimproved sanitation facility** | 1.07 | 0.82, 1.38 | 0.626 | 1.02 | 0.7, 1.48 | 0.926 | 1.27 | 1.05, 1.53 | 0.013 |

Note: adjusted for: age at enrolment, parity, average MUAC <23 cm (Underweight), household located in rural area, education, % likelihood below the poverty line and season at conception.

Abbreviations: CI, confidence interval; aOR, adjusted odds ratio; PRECISE, PREgnancy Care Integrating translational Science, Everywhere; SGA, small-for-gestational-age

**Figure S1.** Correlation matrix of variables with a p-value less than 0.2 in the univariate models

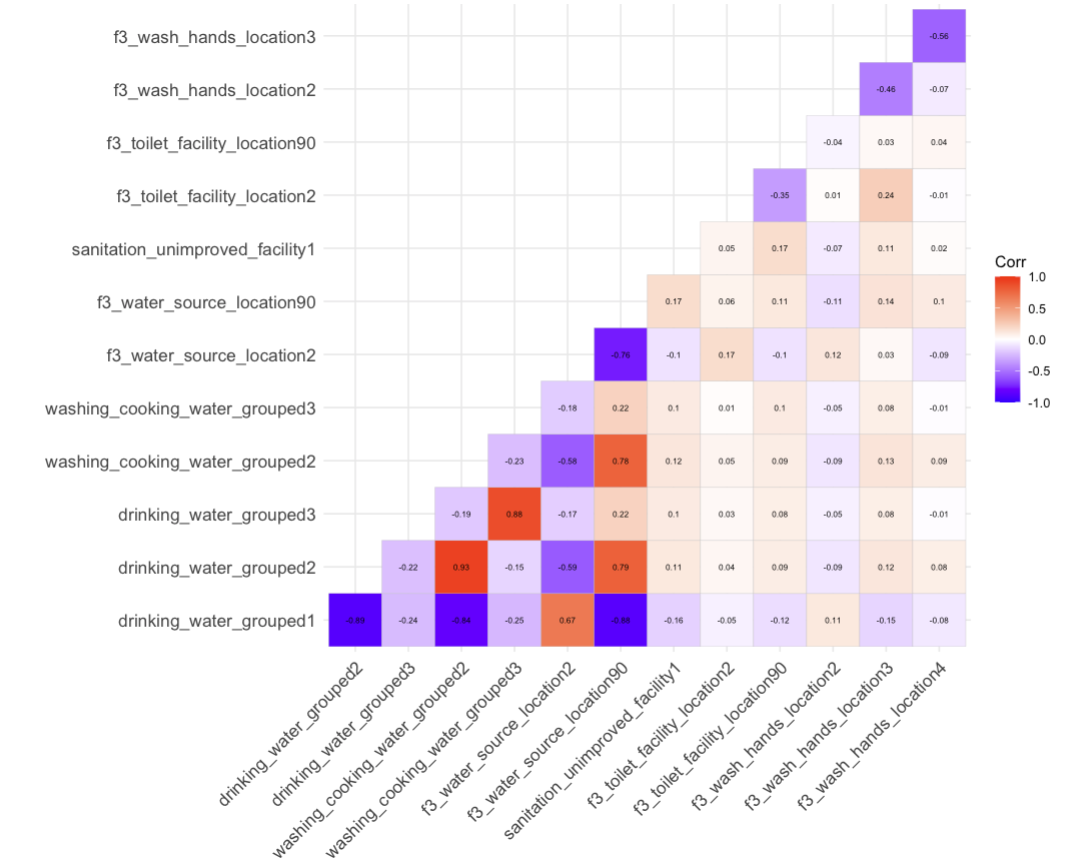
